# Interferon-λ-Neutralizing Autoantibodies and Common Autoimmune Disease Autoantibodies in Pediatric Acute-Onset Neuropsychiatric Syndrome

**DOI:** 10.1101/2024.11.10.24317007

**Authors:** Xihui Yin, Jennifer Frankovich, Muge Kalaycioglu, Ananya Choudhury, Michael J. Burry, Woo Joo Kwon, Meiqian Ma, Bahare Farhadian, Cindy Manko, Melissa Silverman, Yuhuan Xie, Paula Tran, Noelle Schlenk, Paul J. Utz, Tyler R. Prestwood

## Abstract

Pediatric Acute-onset Neuropsychiatric Syndrome (PANS) is characterized by sudden onset obsessive-compulsive disorder (OCD) symptoms in conjunction with other neuropsychiatric manifestations including disturbances in sleep, urination, cognition, and behavior with increased emotional lability and irritability. Clinical observations, including physical exam and laboratory findings, show many similarities between PANS cases and pediatric autoimmune conditions. While published studies have suggested autoantibodies are common in PANS patients, the molecular targets of these autoantibodies (AAbs) remain poorly characterized. Here, we profile the AAbs in 225 plasma samples taken from 166 PANS patients during periods of active disease, or flares, in comparison with 83 pediatric healthy controls using custom Luminex microbead panels conjugated with highly curated antigens from common autoimmune diseases as well as cytokines and chemokines. We find that PANS patients exhibit increased prevalence of AAbs against autoantigens known to be targeted scleroderma and GI/endocrine autoimmune conditions, reflective of clinical observations. Remarkably, a subset of PANS patients exhibits AAbs against interferon-λ, which is an important line of defense against infections at anatomic barriers. Furthermore, 9 of the 11 patients with IFN-λ-binding AAbs were found to be exhibit neutralizing activity. Overall, these findings demonstrate an autoimmune component to PANS and provide insight into a potential disease mechanism mediated by immune deficits at barrier surfaces.

## INTRODUCTION

Obsessive-compulsive disorder (OCD) affects 1-4% of children worldwide and typically exhibits a gradual onset, often beginning in adolescence. In a subset of cases, however, severe OCD symptoms begin abruptly following infections, often streptococcal pharyngitis.^1–7^ Detailed clinical studies have led to the designation by expert panel consensus of Pediatric Autoimmune Neuropsychiatric Disorders Associated with Streptococcal infections (PANDAS) and the broader Pediatric Acute-onset Neuropsychiatric Syndrome (PANS) as subcategories of pediatric OCD.^8,9^ In these cases, a post-infectious process driven by the immune system is strongly suspected.

Clinical evidence supports an autoimmune component of PANS. At presentation, PANS patients may exhibit choreiform movements one of the major Jones criteria for Acute Rheumatic Fever (ARF) diagnosis.^10^ Furthermore, Sydenham chorea is strongly associated with streptococcal infections and the development of anti-streptococcal antibodies that cross-react with central nervous system (CNS) antigens.^11,12^ Common clinical manifestations in PANS overlap with pediatric rheumatological conditions including elevated levels of immune complexes, complement activation (low C4 with normal C4 gene copy number), enthesitis, arthritis, anemia due to inflammation, periungual redness and swelling (a finding in scleroderma and dermatomyositis), livedo reticularis, and other signs suggestive of small vessel inflammation.^13,14^ Additionally, these patients develop additional autoimmune conditions (beyond PANS and arthritis) years after PANS onset including thyroiditis, inflammatory bowel disease, etc.^13^ Furthermore, brain imaging studies, polysomnography studies (indicating movements during REM sleep), and neurological soft signs point towards an process in basal ganglia structures similar to findings in Sydenham chorea and other basal ganglia disorders (i.e. Parkinson’s).^10,15,16^ Although many PANS patients have laboratory evidence of autoimmunity and inflammation, including presence of antinuclear antibodies (ANA), molecular targets remain unknown. Indeed, increased antibody binding to mouse brain tissue has been observed using tissue sections incubated with PANS sera, particularly in brain regions implicated in OCD.^17,18^ However, targets of the AAbs in PANS remain poorly characterized.

Increasing evidence points to infections as triggers for autoimmunity and immune dysfunction. Indeed, patients infected with SARS-CoV-2 or other viruses develop new-onset autoantibodies (AAbs) including anti-cytokine antibodies (ACA) with blocking activity, and multiple large epidemiologic studies have shown significantly increased prevalence of new AID in SARS-CoV-2- infected patients^19–22^. Moreover, high-profile reports implicate Epstein Barr Virus (EBV) in multiple sclerosis (MS) and systemic lupus erythematosus (SLE).^23–25^ Collectively, these studies strongly suggest that bacterial and viral infections can trigger AID and CNS disease.

The relationship between infections and autoimmunity is bidirectional--in addition to infections triggering autoimmunity, self-directed antibody responses are also known to impact the course of infections. For example, pre-existing, type I interferon (IFN)-neutralizing AAbs have been associated with severe COVID-19, resulting in an estimated 20% of COVID-19-related deaths.^26,27^ Type I IFN-neutralizing AAbs have also been associated with exacerbated viral encephalitis, and influenza and herpesvirus infections, as well as an increased likelihood of recurrent infections.^20,28–32^ Overall, these studies point to cytokine-neutralizing AAbs as determinants of disease severity.

Given this evidence, we hypothesized that AAbs may contribute to disease in PANS. Here, we profile AAbs using plasma samples collected from a large cohort of patients with PANS taken during active disease compared with healthy controls (HC). We utilize custom Luminex microbead-based panels consisting of highly curated autoantigens from common AID, as well as cytokines, chemokines and growth factors. Overall, at a population level AAb profiles between PANS patients and HC demonstrate an increased prevalence of AAb particularly against autoantigens related to scleroderma/dermatomyositis and GI/endocrine autoimmunity, reflecting clinical disease observations. Furthermore, we also identify novel IFN-λ-binding AAbs in a subset of PANS patients.

Most of these IFN-λ AAbs are potently neutralizing, which is known to be rare in the general population.^33^ Overall, these data provide further support for autoimmunity in PANS and identify a novel molecular pathway of potential importance in inflammatory CNS disease.

## MATERIALS AND METHODS

### Study Cohorts

#### Selection criteria

The Stanford Immune Behavioral Health (IBH)/PANS clinic evaluated 458 consecutive patients who met clinic entry criteria between September 15, 2012 and January 6, 2023. Per protocol, all patients accepted for evaluation at the PANS/IBH Clinic were referred by their primary care provider, pre-screened to increase the probability of a PANS diagnosis, have new-onset PANS and reside within the counties surrounding the clinic. The pre-screening process includes a thorough review of medical records and a pre-screening questionnaire; in unclear cases, the case is discussed with the child’s primary doctor and other consultants who have been involved with the case. Psychiatric diagnoses were classified by experienced pediatric psychiatrist (MT, MS, PT, YX). Of 458 patients considered for this study, 225 patients met PANS criteria, and 166 of these patients had at least one sample available in our biobank (in flare) and met criteria below (see Supplemental Figure 1).^8^ Several blood samples were collected from each patient (flare and recovery). Samples were excluded if the patient received IVIG (within 9 months) or Rituximab treatment (within 2 years) prior to collection. Of the patients considered for this study, 166 subjects had at least one sample collected during a flare and were therefore included in this study. An additional flare sample was included if collected more than one year after the initial flare sample (maximum 2 in-flare samples per individual). The samples included in this study cohort (Illuminate 1a) will be included in further research studies (epitope mapping, further autoantibody arrays, proteomics, etc). Pediatric healthy control samples (n=83) were recruited from the same local community and met the following criteria: no known history of autoimmune or psychiatric condition in volunteer or first-degree family member.

#### Plasma samples

Plasma samples from PANS patients were collected between September 19, 2013, and November 17, 2023. Aliquoted plasma samples were stored in the Stanford Biobank. Demographic and clinical information for these samples is described in detail below and in Tables 1-4. Three patients had onset of PANS after a SARS-CoV2 infection (unpublished data).

### Prototype autoantibody control samples

Positive control plasma samples for array experiments were derived from donors with prototype autoimmune disorders with known reactivity (topoisomerase 1 [Scl-70], centromere, Sjögren’s Syndrome type A [SSA], whole histones, mitochondria, thyroglobulin, and ribonucleoprotein [RNP]) and were purchased from ImmunoVision. Serum from patients with Autoimmune Polyglandular Syndrome Type 1 patients were generously provided by Dr. David Lewis (Stanford) and used for array experiments.

### Bead-based autoantigen arrays

Autoantigen arrays were generated and utilized as previously described.^19^ Briefly, to construct the array, 1 × 10^6^ carboxylated magnetic beads per analyte (MagPlex-C, Luminex) were distributed into 96-well plates (Greiner BioOne), washed, and resuspended in phosphate buffer (0.1 M NaH_2_PO_4_; pH 6.2) using a plate magnet. Bead surfaces were activated by addition of 0.5 mg of 1-ethyl-3(3- dimethylamino-propyl)carbodiimide (Pierce) and 0.5 mg of N-hydroxysuccinimide (Pierce) according to Luminex protocol. After 20 minutes of activation, beads were washed into coupling buffer (0.05 M MES (2-(N-morpholino)ethanesulfonic acid); pH 5.0) and incubated with 8 μg of each analyte or control antibodies for 2 hours at room temperature. Conjugated beads were washed and resuspended in storage buffer [0.02% Tween 20 in phosphate-buffered saline (PBS-T), 0.1% BSA, and 0.05% sodium azide], and stored at −80°C. Immobilization of the antigens was confirmed using the prototypical control human plasma samples with known reactivity as described above (Immunovision) or antibodies specific for 6× histidine epitope tags (Abcam, ab9108).

For array probing, plasma samples were diluted 1:100 in an assay buffer of 0.05% PBS-T supplemented with 1% (w/v) bovine serum albumin (BSA, Sigma-Aldrich) in a 96-well plates. The bead array was distributed into a 384-well plate (Greiner) by transfer of 5 μL of bead array mixture per well, and then 45 μL of the diluted sera were added into the 384-well plate. Samples were incubated for 60 min on a shaker at room temperature. Beads were washed three times with 60 μL of PBS-T on a plate washer (EL406, BioTek). Beads were resuspended in 50 μL of 1:1000 diluted R-phycoerythrin (PE)–conjugated Fcγ fragment–specific goat anti-human IgG (109-116-098, Jackson ImmunoResearch) in 0.05% PBS-T and incubated at room temperature for 30 min. Plates were washed three times with 60 μL of PBS-T and resuspended in 50 μL of PBS-T for analysis using a FlexMap3D instrument (Luminex Corporation). Binding events were displayed as Median Fluorescence Intensity (MFI). Duplicate plates were run for accuracy and quality control.

### IFN functional assays

IFN-λ functional assays were conducted using IFN-λ Reporter HEK 293 (HEK-Blue IFN-λ) cells (InvivoGen, hkb-ifnlv2). The commercial cell line, which is specific for type III IFN stimulation, contains stable overexpression of the human IFN-λ receptor (interferon lambda receptor 1 (IFNLR1) and interleukin-10 receptor 2 (IL10R2) chains) and secreted embryonic alkaline phosphatase (SEAP) under the control of the IFN-stimulated response element (ISRE). HEK-Blue IFN-λ cells were maintained and subcultured in growth medium supplemented with selective antibiotics following manufacturer’s instructions. For functional assays, HEK-Blue IFN-λ cells were seeded with IFN-λ into half-area 96-well plates at a density of 1.12 × 10^4^ cells per well without selection antibiotics and incubated for 24 hours. Each IFN-λ isoform was titrated using 3.16-fold serial dilutions. SEAP activity was measured with the data fitted to a Four Parameter Logistic (4PL) Curve using GraphPad Prism. The excitatory concentration required for 75% IFN-λ stimulation (EC75) was calculated using the following equation:

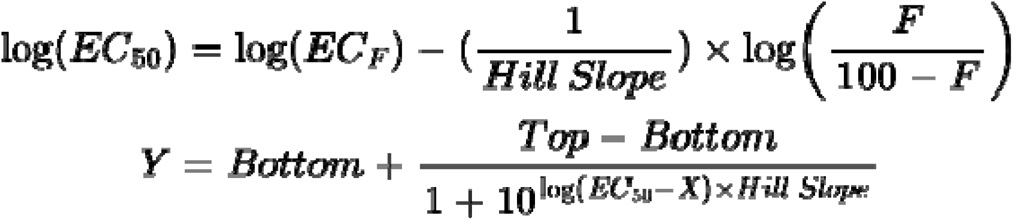

where:

“Top” and “Bottom” are the maximum and minimum plateau values of the Y axis, respectively “Hill Slope” describes the steepness of the curve

EC50 is the concentration of IFN-λ required to reach a SEAP readout halfway between the “Top” and “Bottom” values

ECF is the concentration of IFN-λ required to achieve F% of the SEAP readout between “Top” and “Bottom” values, where F = 75.

To validate the IFN-λ neutralization assays, anti-IFN-λ monoclonal antibodies specific for each IFN-λ isoform were serially diluted 10-fold in growth medium and incubated for 30 minutes with the respective IFN-λ isoforms at their EC75 concentrations (IFN-λ1 = 0.5 ng/mL, IFN-λ2 = 1.1 ng/mL, IFN-λ3 = 1.4 ng/mL) before adding to cells (Figures 2 and S2). To assess the neutralization activity of plasma, samples were heat-inactivated at 56°C for 30 minutes. Plasma from patients or healthy control was diluted to 10% final concentration incubated for 30 minutes with IFN-λ at its EC75 concentration before addition to the freshly seeded HEK-Blue IFN-λ cells. Alternatively, purified plasma IgG and the corresponding IgG-depleted flow-through were diluted to match the IgG or plasma concentrations of the original samples. After 24 hours of stimulation, 20□µL of supernatant from each well was mixed with 180□µL of QUANTI-Blue™ solution (InvivoGen, rep-qbs) and incubated for 45 minutes. SEAP levels were measured by absorbance at 620 nm (OD 620) on a microplate reader (Biotek Synergy HT). Relative IFN-λ levels were determined by SEAP activity interpolated using a reference IFN-λ activity titration curve on the same plate. Activity values were normalized to control wells containing purified IFN-λ. The IFNLR1 neutralization assays were modified so that 10% plasma from patients or healthy controls was directly incubated with HEK-Blue IFN-λ cells, rather than with IFN-λ as above, for an hour before stimulation with IFN-λ1.

The vendors and catalog numbers for the monoclonal antibodies against IFN-λ isoforms and recombinant IFN-λ proteins are listed in Supplemental Table 1.

### IgG purification

A volume of 20 µL protein G magnetic resin (Lytic Solutions) was added to wells of a PCR plate and then washed with 100 µL PBS using a plate magnet. Resin was resuspended in 50 µL plasma diluted 1:1 with PBS. The plasma-resin mixture was incubated for 3h to overnight at 4°C on rotating mixer with inversion to maintain resin suspension. Samples were collected and subjected to subsequent rounds of incubation with magnetic resin until IgG levels became undetectable, as determined by ELISA described below. Resin with IgG bound was washed five times with 200 µL PBS and resuspended in 44 µL 100 mM glycine pH 2.4. This was incubated for 5 minutes at room temperature with periodic resuspension to elute IgG. The purified IgG in the supernatant fraction was collected, then 6 µL 1M Tris pH 8.0 was added to neutralize the solution.

### Quantification of human total IgG by ELISA

Human IgG concentrations were quantified using a human IgG ELISA Kit (Invitrogen™ 885055022) following the manufacturer’s instructions.

### Data processing and statistical analyses

Autoantigen array data processing, quality control and heatmap generation was carried out using R (4.2.3) and RStudio (RStudio Team, 2023.12.1). For quality control, wells with bead counts below 25 for any given antigen(s) were flagged. If the coefficient of variation (CV) of the replicate antigen MFI values was greater than 20% in the flagged well, the MFI value for that antigen was removed from analyses. Additionally, wells with positive-control antigen MFI values (human IgG from serum, anti-Human IgG Fc-fragment specific, anti-human IgG (H+L), anti-human IgG F(ab’) fragment-specific) that were below or above 2.5 standard deviation (SD) from the mean were also discarded from analysis. Finally, MFI values for “bare bead” IDs were subtracted from MFI values for antigen conjugated bead IDs, with MFI averages calculated after background subtraction. ggplot2 (3.5.0) was used for heatmap generation.

For data visualization and statistical analyses, processed datasets were imported into GraphPad Prism (10.3.1) (cite). Statistical differences in MFI values between patients and healthy controls for each antigen were determined using two-tailed Mann-Whitney U tests with Benjamini-Hochberg correction (FDR < 0.05) or using Fisher’s exact test for general autoantibody prevalence using a threshold of 5 SDs above the mean HC median fluorescence intensity (MFI) and a 3,000 MFI static cutoff. P values less than 0.05 were considered significant.

### Data and code availability

Deidentified array data is publicly available on the Gene Expression Omnibus (GEO) database. The accession code is provided in our data availability statement.

## RESULTS

### Demographic parameters of Illuminate 1 Cohort

The course of PANS is characterized by a relapsing-remitting pattern, marked by flares (sudden exacerbations of psychiatric symptoms), followed by intervals of relative symptom stability or remission.^34,35^ Patients (n=166, flare draws= 225) had a median age (at first flare sample collection) of 11.3 years (x□=11.5, s=4.3, range 4-26). Controls (n=83) had a median age (at sample collection) of 13.7 years (x□=14.3, s=5.0, range 4- 24). Demographics of our patient and control cohorts are shown in **Table 2**.

**Table 1.** Count of blood specimens by type (flare v. recovery) from PANS subjects from the Illuminate 1 cohort.

| Count of blood specimens by type (flare v. recovery)<br>(Stanford Illuminate 1 Cohort) | PANS (n=166) |
| --- | --- |
| First flare research draw captured in clinic | 166 (100%) |
| Second flare research draw captured in clinic<br>(>1 year after first flare) | 59 (36%) |
| Best recovery research draw | 98 (59%) |

**Table 2.** Demographics of PANS subjects from the Illuminate 1 cohort and healthy controls.

| **Demographics of PANS cohort and healthy controls**<br>**(Stanford Illuminate 1 Cohort)** | **PANS (n=166)** | **Control (n=83)** |
| --- | --- | --- |
| Age at disease onset, years (mean) | 9.5 ± 3.7 | - |
| Age at clinic presentation, years (mean) | 10.6 ± 3.9 | - |
| Age at first flare blood draw, years (mean) | 11.5 ± 4.3 | 14.3 ± 5.0 |
| Sex |  |  |
| Male | 101 (61%) | 41 (49%) |
| Race |  |  |
| White | 138 (83%) | 43 (52%) |
| Asian | 5 (3%) | 29 (35%) |
| Two or more races reported | 22 (13%) | 10 (12%) |
| Other | 1 (<1%) | 1 (1%) |
| Ethnicity |  |  |
| Hispanic or Latino | 23 (14%) | 12 (10%) |

Note: Data are presented in number (percentage) or mean ± SD unless specified.

### PANS patients show clinical signs of autoimmunity

As part of clinical evaluation and treatment, patients undergo detailed clinical examination and laboratory workup. Clinical examination commonly demonstrates stigmata of autoimmune disease including arthritis, enthesitis, high levels of immune complexes, low C4, periungual redness and swelling, etc (**Table 4**).

**Table 3.** Symptoms at clinical presentation in PANS subjects from the Illuminate 1 cohort. **Abbreviations:** OCD, obsessive compulsive disorder.

| Symptoms at Clinic Presentation (Stanford Illuminate 1 Cohort) | PANS (n=166) |
| --- | --- |
| Neuropsychiatric symptoms at clinic presentation, n (%) |  |
| OCD | 146 (88%) |
| Eating restriction (food refusal or avoidance) | 82 (49%) |
| Anxiety | 149 (90%) |
| Emotional lability and/or depression | 144 (87%) |
| Irritability, aggression and/or severely oppositional behaviors | 136 (82%) |
| Behavioral/developmental regression | 76 (46%) |
| Cognitive impairment | 85 (51%) |
| Sensory amplification | 97 (58%) |
| Motor issues | 63 (38%) |
| Urinary symptoms | 70 (42%) |
| Sleep disturbances | 113 (68%) |
| Flare Status at Clinic Presentation |  |
| In flare | 136 (82%) |
| Not in flare | 20 (12%) |
| Unclear flare / No data | 10 (6%) |
| Psychometric scores at (or nearest) clinic presentation |  |
| PANS Global Impairment Score (GIS) Range (0-100)<sup>A,B</sup> | 47.9 ± 23.4 |
| Caregiver Burden Inventory (CBI) Range (0-96)<sup>A,C</sup> | 34.2 ± 18.0 |
| Caregiver Burden Inventory (CBI) ≥ 36<sup>C,D</sup> | 75 (47%) |

Note: Data are presented in number (percentage) or mean ± standard deviation
<sup>A</sup>Higher scores indicate more impairment and burden for GIS and CBI
<sup>B</sup>Of patients with available data (n=165)
<sup>C</sup>Of patients with available data (n=161)
<sup>D</sup>36 is the threshold for needing respite care

**Table 4.** Clinical Findings. Arthritis (according to ILAR and ASAS criteria),^53,54^ inflammatory back pain (according to Calin criteria),^55^ autoimmune disease, and markers of inflammation at clinic presentation. **Abbreviations:** ASAS, Assessment of SpondyloArthritis international Society; ILAR, International League of Associations for Rheumatology.

| Clinical findings | PANS (n=166) |
| --- | --- |
| Any arthritis (meets ILAR and/or ASAS criteria) | 50 (30%) |
| Arthritis subtype (criteria met) | 33 (20%) |
| Enthesitis-related arthritis (ILAR) | 10 (6%) |
| Psoriatic arthritis (ILAR) | 2 (1%) |
| Oligoarticular arthritis (ILAR) | 25 (15%) |
| Spondyloarthritis (ASAS) | 43 (26%) |
| Inflammatory back pain (Calin criteria) | 24 (14%) |
| Any autoimmune/inflammatory disease (beyond arthritis)<sup>A,B</sup> | 99/153 (65%) |
| Antinuclear antibody, titer $\geq 1:80$ | 22/99 (22%) |
| Antihistone antibody, high | 16/70 (23%) |
| Antithyroid antibody high<sup>C</sup> | 21/112 (19%) |
| C1q binding assay, high | 25/120 (21%) |
| Complement 3, low | 11/124 (9%) |
| Complement 4, low | 41/126 (33%) |
| von Willebrand factor antigen, high | 14/116 (12%) |
| D-dimer, high | 6/111 (5%) |
| Physical examination findings (any)<sup>D</sup> | 74/132 (56%) |
| Livedo reticularis, present | 60/84 (71%) |
| Periungual redness, present | 30/42 (71%) |
| Onychodermal band, abnormally prominent | 54/79 (68%) |

Note: Data are presented in number (percentage) or mean ± SD unless specified.
<sup>A</sup>Some patients carry more than one diagnosis
<sup>B</sup>These are thyroiditis (n=9), psoriasis (n=7), Celiac disease (n=3), chronic urticaria (n=3), primary immunodeficiency (n=2), inflammatory bowel disease (n=2), eosinophilic esophagitis (n=2), systemic lupus erythematosus (n=1), diabetes mellitus (n=1), period fever (n=1)
<sup>C</sup>Thyroglobulin antibodies and/or thyroid peroxidase antibodies
<sup>D</sup>All the laboratory and physical examination signs presented were hypothesized to be relevant in the fall of 2012, and the clinicians have attempted to collect the data at the first flare captured in the clinic for all patients. Due to the severe psychiatric symptoms and young age of the study sample, we were not able to complete the standard workup for all patients; denominators are reported for each marker.

### A subset of the PANS Illuminate 1 Cohort exhibit high levels of AAbs associated with scleroderma and GI/endocrine autoimmunity

Approximately 1/4 of PANS patients exhibit anti-nuclear antibodies (ANA). Despite these observations, the autoantibody repertoire in PANS has not been characterized in detail. Therefore, we sought to evaluate the prevalence of AAbs characteristic of traditional autoimmune disease during PANS flares. To assess the breadth of AAbs targeting common autoantigens in connective tissue disease (CTD), we implemented a custom 28- plex traditional autoantigen array made up of highly curated antigens validated through previous studies.^19,20^ The autoimmune diseases represented by these antigen panels include systemic lupus erythematosus, Sjögren syndrome, dermatomyositis, scleroderma, primary biliary cirrhosis, thyroiditis, pernicious anemia, and autoimmune vasculitides. A panel of prototype sera with known reactivity to several of these antigens served as positive controls.

Using a threshold of 5 SDs above the mean HC median fluorescence intensity (MFI) and a 3,000 MFI static cutoff, we assessed the prevalence of AAbs to these targets (**Fig 1A**). Three samples were excluded from analysis due to technical failure or disease reclassification. We observed several “hits” in the PANS group with signal often far exceeding that of HC. The prevalence of AAbs was elevated in PANS patients relative to HC as determined by Fisher’s exact test specifically for the scleroderma and GI/endocrine panels, (p=0.0089 and p=0.0161). More specifically, the autoantigens targeted in PANS patients from the scleroderma panel included CENP A (Centromere Protein A), Fibrillarin (U3 small nucleolar ribonucleoprotein), Scl-70 (topoisomerase I) and U11/12 RNP (ribonucleoprotein), and from the GI/endocrine panel included PDC-E2 (pyruvate dehydrogenase complex E2, a mitochondrial antigen targeted in primary biliary cirrhosis), thyroid antigens, and intrinsic factor (pernicious anemia) (**Fig 1B**). Additionally, while the prevalence across the myositis/overlap syndrome panel did not differ statistically between PANS flares and HC (p = 0.6171), several autoantigens including MDA5 (an antiviral, RNA-binding self-protein targeted in myositis), OJ (a tRNA synthetase target in myositis), and PM-Scl75 (an exosome protein autoantigen in overlap syndromes) exhibited a trend towards increased AAb prevalence and MFI values in PANS flare samples compared with HC. Of note, PANS flare samples did not exhibit an increased level of autoantibody binding against β-tubulin or GM1 ganglioside, previously described targets in Sydenham chorea (**Supp Fig 1**). Overall, the differences in AAb levels observed in PANS were due to an increased prevalence of AAbs in the specific AID subpanels described above, rather than group differences in mean AAbs levels, which were not statistically significant.

**Figure 1.**
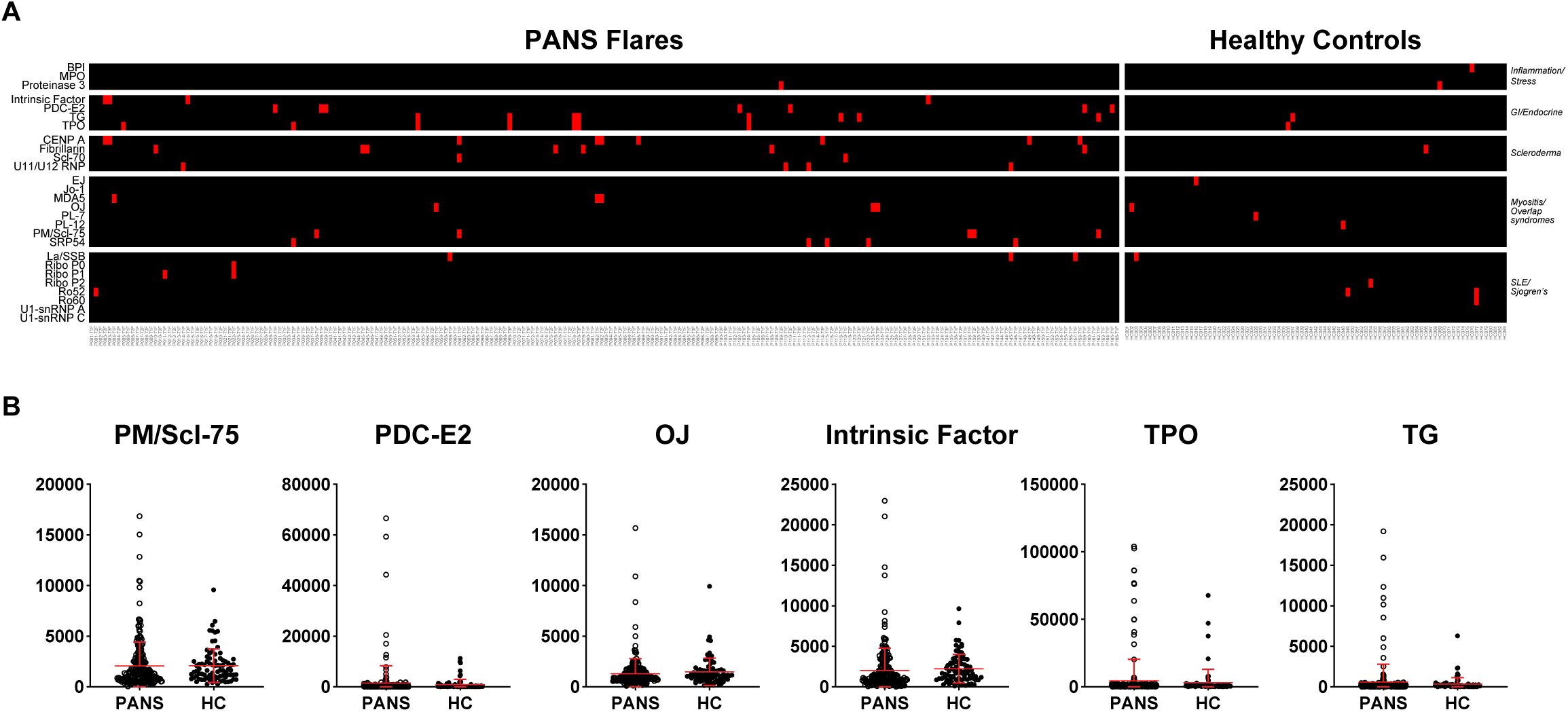
**A)** Heatmap of the IgG autoantibodies discovered using a 28-plex bead-based protein array. Autoantigens are grouped based on associations with autoimmune diseases, tissue inflammation or stress responses. Samples screened include PANS flares (left, n = 225), healthy controls (HC, right, n = 83). Positive values are represented in red using a threshold of 5 SDs above the mean HC MFI and a 3,000 MFI static cutoff. **B)** Violin dot plots of specific antigens from the scleroderma and GI/endocrine panels showing outliers with positive signal. Middle red lines denote to the median MFI, while the lower and upper red lines correspond to the first and third quartiles, respectively. Statistical significance for relative prevalence of AAbs against each subpanel was determined with Fisher’s exact test comparing PANS flare and HC groups (P < 0.05).

### A subset of patients with PANS produce AAbs that bind to type I and type III interferons

Anti-cytokine AAbs (ACA) are increasingly recognized as important modulators of disease induced by infections, most recently with COVID-19 disease severity and anti-type I interferon AAbs.^19,26,36^ Given the observed post-infectious nature of PANS, we characterized the ACA repertoire in PANS patients during flares. To do this, we used the samples described above collected during PANS disease flares and compared samples from HC using a custom 56-plex panel composed of 12 interferon molecules, 14 interleukins, 18 chemokines, and 12 other cytokines. Several individual patients exhibited high levels of AAbs against type I and type III IFNs, though, as above, we did not detect group differences in mean AAb levels overall (**Fig 2A**). More specifically, we detected outlier patients with AAbs against IFN-α7, IFN-α8, IFN-α10, and IFN-β, as well as a larger, non-overlapping group producing AAbs against IFN-λ1, IFN-λ2 and IFN-λ3 (**Fig 2B**).

**Figure 2.**
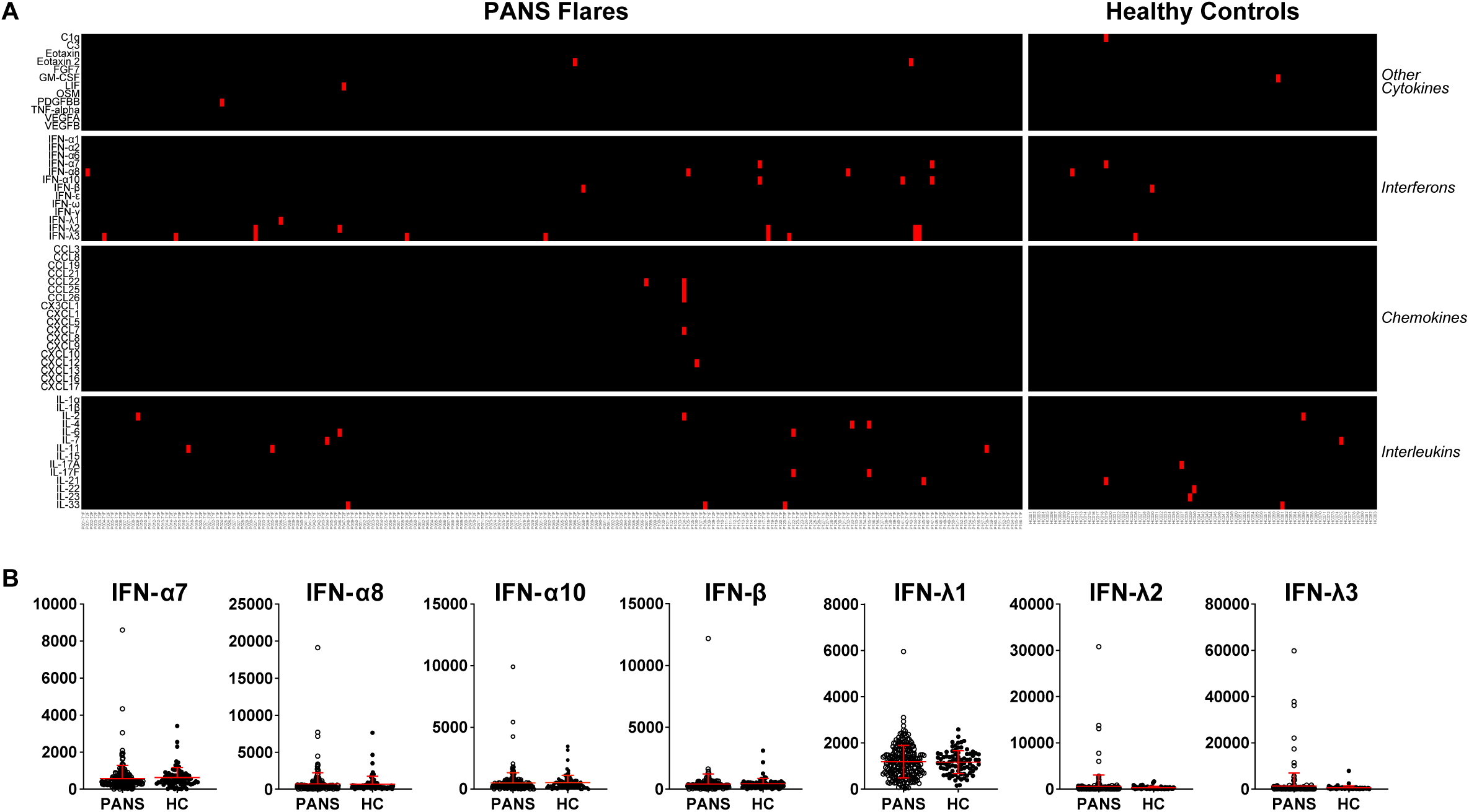
**A)** Heatmap of the IgG autoantibodies discovered using a 56-plex bead-based protein array. Autoantigens are grouped based on class of immune molecules including IFNs, chemokines, interleukins and other cytokines. Samples screened include PANS flares (left, n = 225) and HC (right, n = 83). Positive values are represented in red using a threshold of 5 SDs above the mean HC MFI and a 3,000 MFI static cutoff. **B)** Violin dot plots of specific antigens from the IFN panels showing many of the outliers with positive signal. Middle red lines denote to the median MFI, while the lower and upper red lines correspond to the first and third quartiles, respectively. Statistical significance for relative prevalence of AAbs against each subpanel was determined with Fisher’s exact test comparing PANS flare and HC groups (P < 0.05).

### Developing SEAP-based IFN neutralization assays

To assess the functional effects of the identified IFN-neutralizing AAbs, we developed assays utilizing HEK Blue reporter cell lines (InvivoGen, San Diego, CA, USA), which produce secreted embryonic alkaline phosphatase (SEAP) in response to stimulation with specific cytokines (schematic shown in **Fig 3A**). Using these cells, we first validated their capacity to detect IFNs over a range of concentrations (**Fig 3B**). Next, we assessed the ability of this assay to detect the presence of neutralizing antibodies. Using a fixed concentration of IFN at approximately the EC75 (75% maximal effective concentration), we performed titrations of purified, neutralizing monoclonal antibodies against each IFN species. For example, increasing concentrations of IFN-λ3 displayed on a logarithm transformed scale show a sigmoidal response of SEAP signal secreted by the HEK-Blue IFN-λ-reporter cells (**Fig 3B**, lighter line). A reverse sigmoid response is observed when the concentration of IFN-λ3 is held constant at the EC75 of 1.4 ng/mL and increasing levels of IFN-λ3-neutralizing antibody are incubated with the cytokine prior to exposure to the target cells (**Fig 3B**, darker line). Very similar results were obtained with IFN-λ1 with EC75 of 0.5 ng/mL and IFN-λ2 with EC75 of 1.1 ng/mL (**Supp Fig 2**).

**Figure 3.**
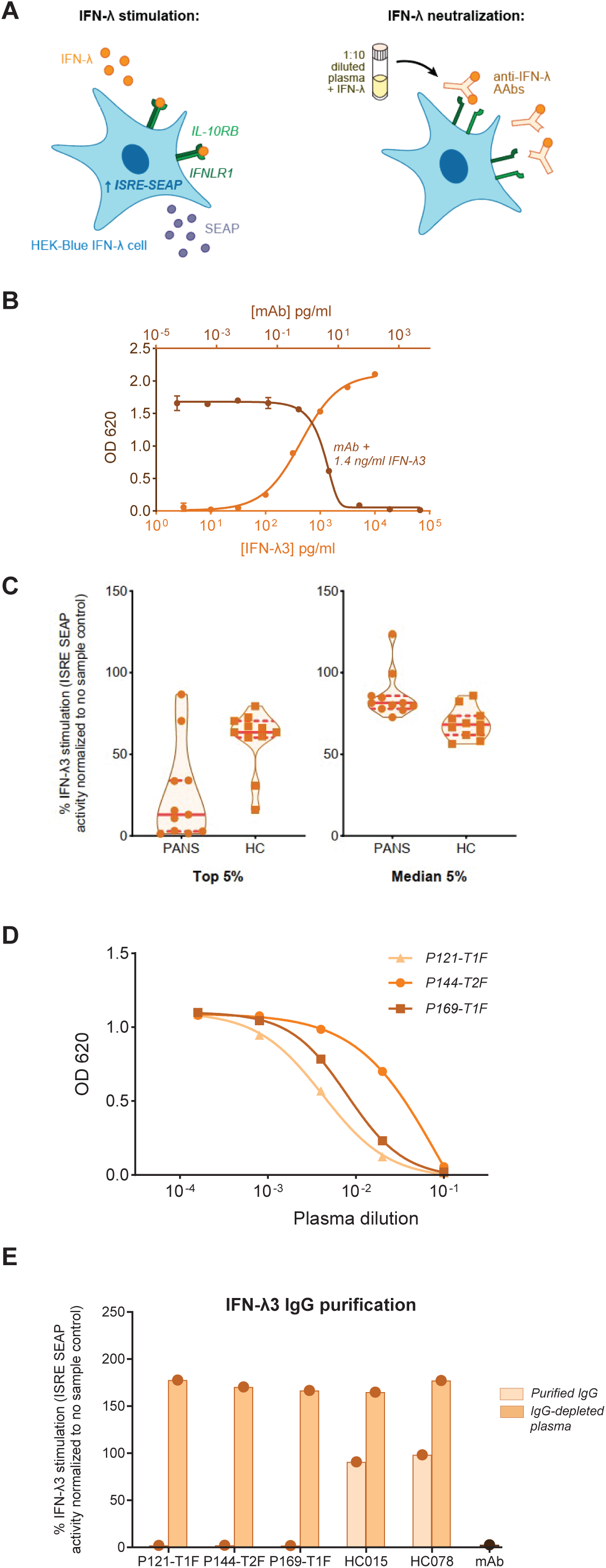
**A)** Schematic overview of the IFN-λ neutralization assay using HEK-Blue IFN-λ cells. Cells contain an ISRE-SEAP reporter system specific for Type III IFN stimulation. IFN-λ is incubated with 10% plasma before addition to cells. **B)** Validation of the ISRE-SEAP reporter system in HEK-Blue IFN-λ cells with serial dilutions of IFN-λ3. Also, dose-dependent neutralization of IFN-λ3 at the EC_75_ concentration of 1.4 ng/mL in the presence of serially diluted anti-IFN-λ3 antibody (5 x 10^-4^ to 5 x 10^3^ ng/mL) mAbs reflect sensitivity of the neutralization assay. **C)** IFN-λ3 neutralization assay results comparing patients and healthy control groups. Samples were selected based on the top or median 5% (n = 11) of MFI values from the IFN-λ autoantibody array screen (from **Fig 2A**). Values reflect relative IFN-λ3 stimulation (at the EC_75_ concentration of 1.4 ng/mL) in the presence of a 1:10 dilution of heat-inactivated plasma after normalization to IFN-λ-only control. **D)** Neutralizing activity of patient plasma with fully neutralizing IFN-λ3 AAbs. Samples were serially diluted 5-fold prior to incubation with IFN-λ3 (1.4 ng/ml). **E)** IFN-λ3 neutralization assays using purified IgG from samples with high neutralizing activity compared with IgG-depleted plasma from the same samples and HC. IFN-λ3 mAb (5 µg/mL) was included as a control. Samples were incubated with IFN-λ3 (1.4 ng/mL).

### PANS patients exhibit neutralizing AAbs against IFN-λ

To determine whether the observed IFN-λ-binding AAbs in the PANS patient flare samples were functionally neutralizing, we assessed the neutralization potential of the top 5% of PANS flares samples (n=11) based on MFI (**Fig 2A**) as well as the top 11 HC samples for comparison. At a final plasma concentration of 10%, 9 of 11 PANS flare samples showed significant neutralization of IFN-λ3, with near-complete neutralization in 4. No neutralization was detected in 2 samples (**Fig 3C**). Interestingly, 2 of the highest-binding healthy control samples also showed moderate neutralization potential. Next, we determined whether IFN-λ neutralization was common in PANS flare samples, even in the absence of high levels of IFN-λ-binding AAbs. We selected the median 5% of IFN-λ3-binding PANS flare samples (n=11) and compared with the median 11 HC samples. None of these samples showed clear neutralization of IFN-λ3 (**Fig 3C**). Very similar results were obtained with IFN-λ2 (**Supp Fig 3**). With IFN-λ1, however, a single patient exhibited AAb-binding by bead-based assay and these IFN-λ1- binding IgG AAbs were also found to be neutralizing (**Supp Fig 4**).

Next, to further characterize the neutralizing capacity of the PANS samples, the top 3 samples with highest binding to IFN-λ3 identified in **Fig 2A** were serially diluted starting at a top final concentration of 10% in the presence of the EC75 concentration of IFN-λ3. These samples all potently neutralized of IFN-λ3 in 10% plasma, but by 1% neutralization was more variable and almost no neutralization was detectable at 0.1% (**Fig 3D**). Furthermore, to assess whether that the effector molecule mediating cytokine neutralization was IgG, we performed Protein A/G resin to isolate IgG and compared with IgG-depleted plasma in the neutralization assay. These results confirmed that the IgG fraction, but not the IgG-depleted plasma, exhibited the neutralizing activity (**Fig 3E**).

Finally, to examine the neutralization potential against other IFN species in which binding was observed in PANS flare samples, neutralization assays using IFN-α7, IFN-α8, IFN-α10, and IFN-β were performed on the samples identified in **Fig 2A** (data not shown). In each case, no neutralization was observed, demonstrating that PANS is selectively associated with neutralizing AAbs against type III IFN and not other IFN types.

### IFN-binding AAbs are dynamic in PANS patients across time but do not correlate with disease states

To assess whether the levels of IFN-λ-binding AAbs correlated with PANS disease activity, we next analyzed AAb-binding to a panel of IFNs using samples from the patients with IFN-λ-binding collected at additional timepoints including during periods of symptomatic improvement or recovery (**Fig 4A**). We found that levels of AAbs did not clearly correlate with disease state, as several patients exhibited the highest levels of IFN-λ-binding AAbs during disease flares, while others exhibited the highest AAb levels during periods of recovery. Furthermore, analysis across time points showed IFN-λ-binding AAb levels were variable over time in some patients and more consistent in others. Lastly, the subtype specificity of IFN-λ-binding AAbs was variable between patients, with some patients recognizing both IFN-λ2 and IFN-λ3, while others exhibited more selective recognition of only one IFN-λ subtype. Overall, these data demonstrate the temporal variability of AAb levels in these patients and unclear association with disease state.

**Figure 4.**
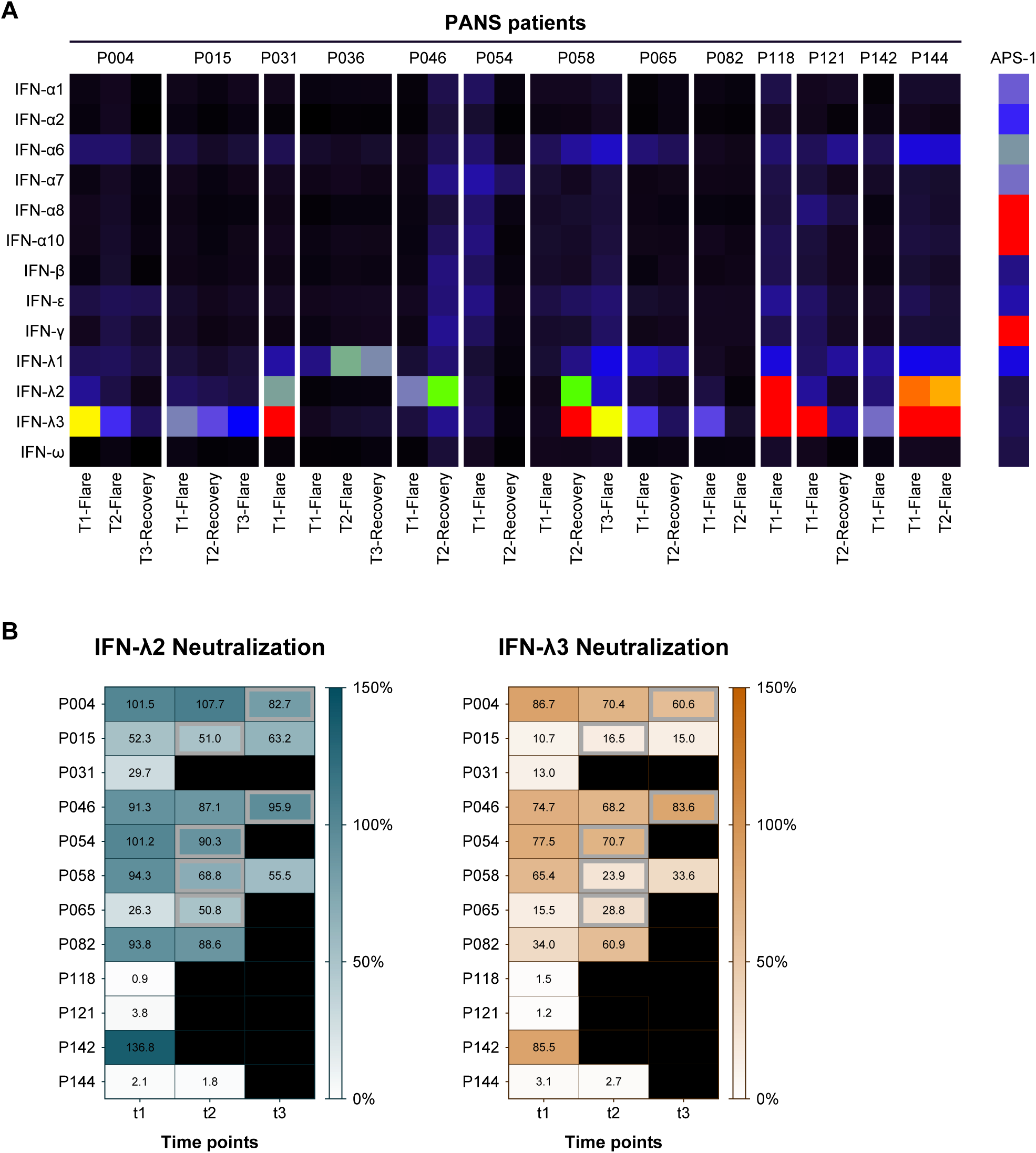
**A)** Heatmap showing anti-interferon IgG autoantibodies discovered using a 13-plex bead-based protein array. The samples include the patients with the top 5% of MFI values for IFN-λ2 and IFN-λ3 from a). Additional patient time points were also included, where available. Autoimmune polyendocrinopathy candidiasis ectodermal dystrophy (APECED, or APS-1) prototype serum and secondary antibody-only wells were included as controls. Colors correspond to the MFI values shown on the right, with values capped at 12,000. **B)** IFN-λ2 or IFN-λ3 neutralization activity of the top 5% of patients for IFN-λ2 and IFN-λ3 reactivity, across available time points. Boxes corresponding to recovery samples are outlined in grey. Values reflect relative IFN-λ stimulation.

For samples with sufficient remaining volume, we examined the neutralization potential at each timepoint (**Fig 4B**). Neutralization capacity of sera was not clearly predictable based on levels of IFN-λ-binding AAbs. Furthermore, some patients exhibited similar levels of AAbs-binding to one IFN-λ subtype, but differing levels of neutralization. Overall, while the subtype specificity of IFN-λ- neutralizing AAbs is heterogeneous, neutralizing capacity is present in most of the patients with IFN-λ-binding AAbs.

Finally, given that multiple patients exhibited neutralizing AAbs against IFN-λ targets, we evaluated the prevalence of AAbs against interferon lambda receptor 1 (IFNLR1), which determines responsiveness to IFN-λ, via Luminex microbead assay with IFNLR1 coupled directly to beads. Similar to the results above, we found that several PANS patients were outliers with much higher binding than HC (**Fig 5A**). We next modified the SEAP-based IFN neutralization assay above to measure effects on cell surface receptors by pre-incubating diluted plasma with cells, prior to exposure to cytokine (schematic shown in **Fig 5B**). Using the samples which exhibited binding only to IFNLR1 (and not to IFN-λ1), we measured the capacity of diluted plasma to block IFN-λ1 activity on target cells across disease states and found that one sample, P006, neutralized IFNLR1 during a disease flare (**Fig 5C**). This result demonstrates that AAbs have the capacity to block signaling not just by binding to the ligand, but also to its cognate cell surface receptor.

**Figure 5.**
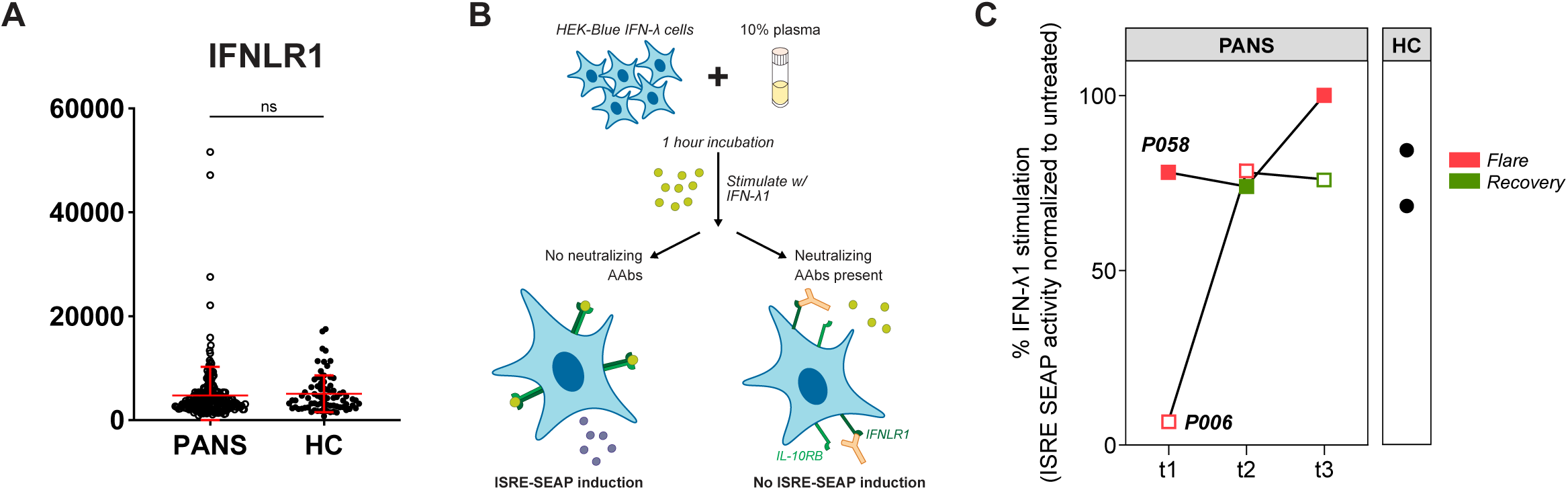
**A)** Violin dot plots of IFNLR1 antibody binding showing outliers with positive signal. Middle red lines denote to the median MFI, while the lower and upper red lines correspond to the first and third quartiles, respectively. **B)** Schematic overview of the IFNLR1 neutralization assay using HEK-Blue IFN-λ cells. Cells contain an ISRE-SEAP reporter system specific for Type III IFN stimulation. Cells are incubated with 10% plasma before addition of IFN-λ1 to cells. **C)** Time course analysis of IFNL1R neutralization activity in two patients across different time points and disease states.

## DISCUSSION

Here, we describe the AAb repertoire against common autoantigens in PANS patients. We found that AAbs associated with scleroderma and GI/endocrine autoimmune diseases have increased prevalence in PANS patients relative to healthy controls. These findings agree with the PANS clinical examination findings of periungal erythema, which is a classic finding associated with scleroderma and dermatomyositis.^37^ The etiologies of scleroderma and dermatomyositis remain unknown, though in both cases distinct subgroups are differentiated by paraneoplastic or postinfectious settings. In scleroderma, a cross-reactive, self-reactive immune response is believed to follow infections including CMV, EBV or Parvovirus B19 or in the setting of environmental exposures.^38–42^ PANS does not have a known neoplastic association, but is associated with infections. Therefore, this data provides further support for PANS as a post-infectious, immune-mediated disease.

We have also discovered a novel association of IFN-λ-neutralizing AAbs with PANS. Type III IFN AAbs were originally described in autoimmune polyendocrinopathy candidiasis ectodermal dystrophy (APECED or APS-1).^43,44^ Since then, few studies have assessed the impact of type III IFN AAbs. Recently, type I IFNs AAbs have been the source of intense investigation recently as they are important determinants of severe disease in COVID-19.^26,27^ Given these findings, the prevalence and function of type III IFN AAbs in the general population and in COVID-19 has also recently been studied.^26,33^ Based on the study by Vanker et al, type III IFN AAbs exhibit a higher prevalence in the general population than type I IFN AAbs (8.5% vs. 3.9%, respectively), yet show a similar increase with age. However, about 2/3 of type III IFN AAbs show no neutralizing activity and there is no known association with severe COVID-19 outcomes.^33^ Accordingly, it has been speculated that type III IFN AAbs may have little functional impact in some contexts, given the redundancies of type I and type III IFN signaling. In contrast to these observations, here, we report a relatively low prevalence of type III IFN AAbs (0.4% in HC) with an increase in PANS patients (2.2%), the majority of whom exhibit neutralization (82%). Furthermore, the detected type III IFN AAb MFIs indicate relatively high levels, in contrast to descriptions of relatively low levels observed in APS-1.^43^ It remains to be determined, however, whether the observed AAbs have relevance to PANS disease.

In contrast to type I IFN, which acts systemically due to the ubiquitous expression of interferon-α/β receptor, IFNLR1, which determines responsiveness to IFN-λ, exhibits expression limited to barrier interfaces.^45^ Studies to date have shown that loss of IFN-λ renders increased susceptibility to infection at barrier surfaces, including the lungs, respiratory tract, gastrointestinal tract, skin and blood-brain barrier.^33,46–49^ In a mouse model of West Nile Virus encephalitis, IFN-λ was found to restrict viral invasion of the CNS by tightening the blood-brain barrier, preventing further dissemination of peripheral molecules into the brain.^50^ These observations may be of particular importance in PANS, where the blood-brain barrier is believed to be compromised in the amygdala and basal ganglia regions.^15,51^ Furthermore, it suggests a potential mechanism in which type III IFN-neutralizing AAbs may prevent IFN-λ-mediated protection of blood-brain barrier during infections or inflammation, rendering the brain more susceptible to invasion by infectious agents or effects of systemic inflammatory mediators.

PANS patients who exhibited IFN-lambda AAbs and other disease-related autoantibodies on our CTD arrays were distinguished by longer time to initial recovery, more frequent flares, longer flares, and higher rate of developing arthritis and/or another autoimmune disease. Two of the PANS patients identified to have IFN-λ-binding AAbs also exhibited relatively rare AAbs against MDA5 and two others had relatively common thyroglobulin-binding AAbs. In dermatomyositis, anti-MDA5 AAbs are associated with the development of rapidly progressive interstitial lung disease.^52^ In these patients, no signs of lung disease were observed. Future studies will aim to assess whether the presence of these type III IFN and other AAbs and other autoantibodies are of clinical significance and whether this finding points toward pathways important in PANS pathogenesis.

It is also important to note that while our observations clearly demonstrate multiple PANS patients with IFN-λ-neutralizing AAbs, this accounts for only a small subset of PANS patients. However, given the relatively low abundance of AAbs in the pediatric population, and the clear association of type I IFN-neutralizing AAbs with increasing age, the occurrence of IFN-λ-neutralizing AAbs in PANS is remarkable. Phenotypically, patients with IFN-λ-neutralizing AAbs were very similar to other PANS patients in terms of symptomatology and disease course, and there were not distinguishing features identified in these patients. Furthermore, for these patients, levels of IFN-λ- neutralizing AAbs were variable over time and across disease states. Therefore, the impact of these AAbs in PANS will require further characterization. The additional observation of a distinct patient with IFNLR1-neutralizing AAbs provides further support for the importance of this pathway in PANS. IFNLR1-neutralization AAbs are remarkable and novel, particularly given that exhaustive investigations that have been unable to identify type I IFN receptor AAbs. Future studies will aim to assess the impact of these IFN-λ-neutralizing AAbs on the blood-brain barrier in PANS, particularly regarding the susceptibility of the brain to infections and inflammation.

## Data Availability

All data produced in the present study are available upon reasonable request to the authors

---

**Supplementary Table 1.**

| Product | Manufacturer | Catalog number |
| --- | --- | --- |
| Interferon-λ1, recombinant human | Peprotech | 300-02L |
| Interferon-λ2, recombinant human | Peprotech | 300-02K |
| Interferon-λ3, recombinant human | PBL Assay Science | 11730-1 |
| Anti-Interferon-λ1 monoclonal antibody | InvivoGen | mabg-hil29-3 |
| Anti-Interferon-λ2 monoclonal antibody | InvivoGen | mabg-hil28a |
| Anti-Interferon-λ3 monoclonal antibody | InvivoGen | mabg-hil28b |

**Supplementary Figure 1.**
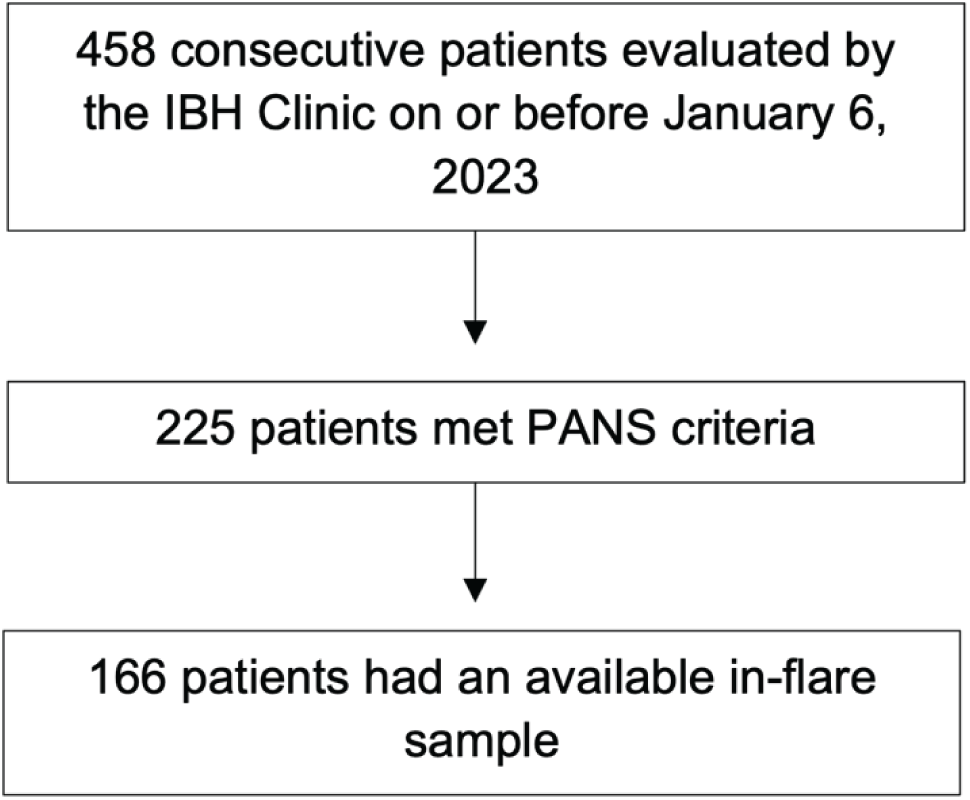
Flowchart of cohort subject selection process.

**Supplementary Figure 2.**
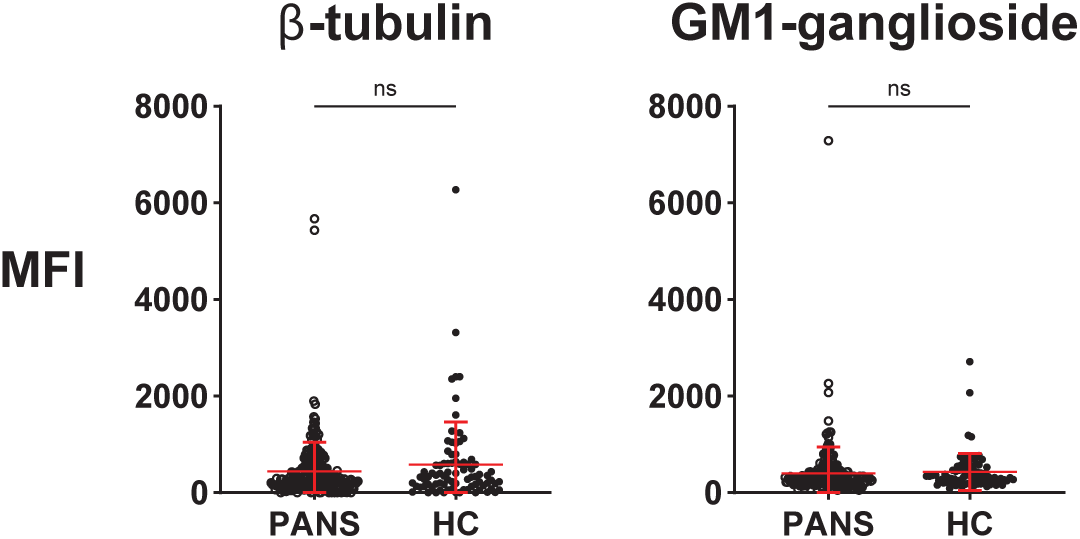
Violin dot plots of antigens associated with Sydenham Chorea (β-tubulin and GM1-ganglioside). Middle red lines correspond to the median MFI, while the lower and upper red lines correspond to the first and third quartiles, respectively. Statistical significance (P < 0.05) was determined between patients and healthy controls groups using two-tailed Mann-Whitney tests with Benjamini-Hochberg correction across all 89 antigens evaluated in this study (FDR ≤ 0.05).

**Supplementary Figure 3.**
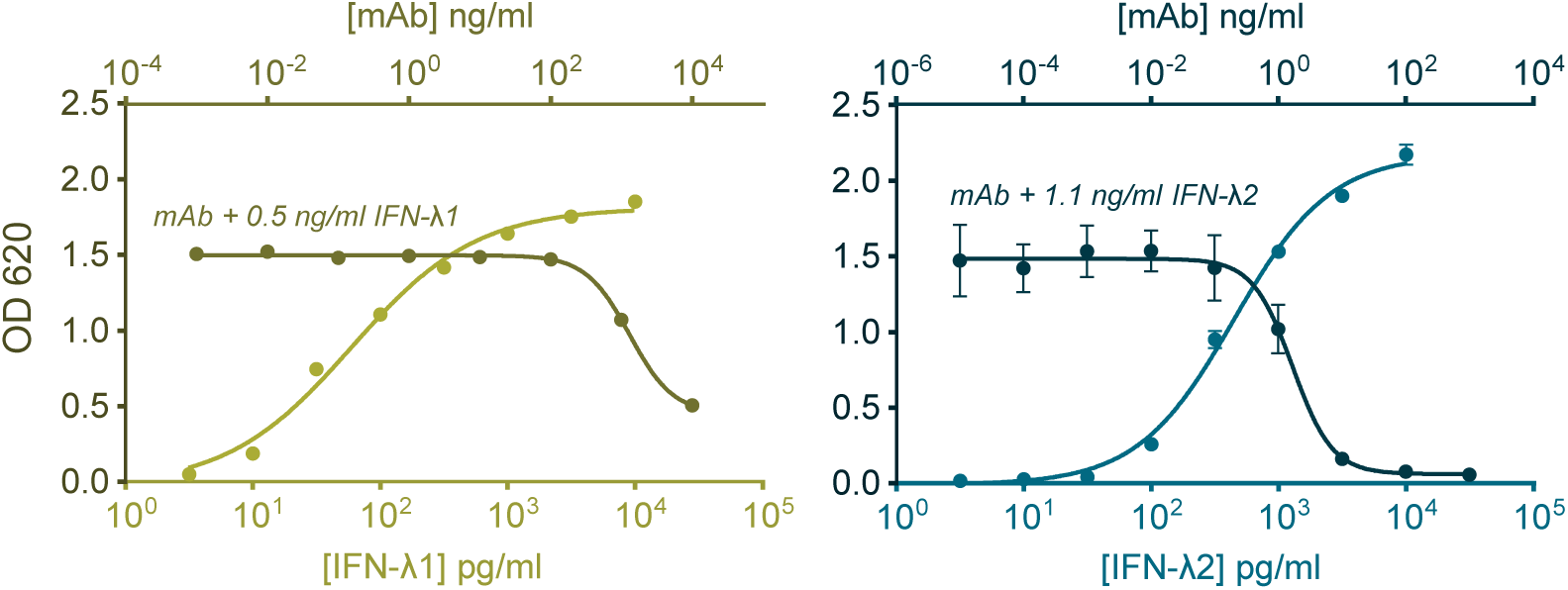
Validation of the ISRE-SEAP reporter system in HEK-Blue IFN-λ cells with serial dilutions of IFN-λ1 or IFN-λ2. Dose-dependent neutralization of IFN-λ1 (0.5 ng/mL) or IFN-λ2 (1.1 ng/mL) at EC_75_ concentrations by anti-IFN-λ1 (10^-3^ to 10^4^ ng/mL) or anti-IFN-λ2 (10^-5^ to 10^3^ ng/mL) mAbs reflect sensitivity of the neutralization assay to various IFN-λ isoforms.

**Supplementary Figure 4.**
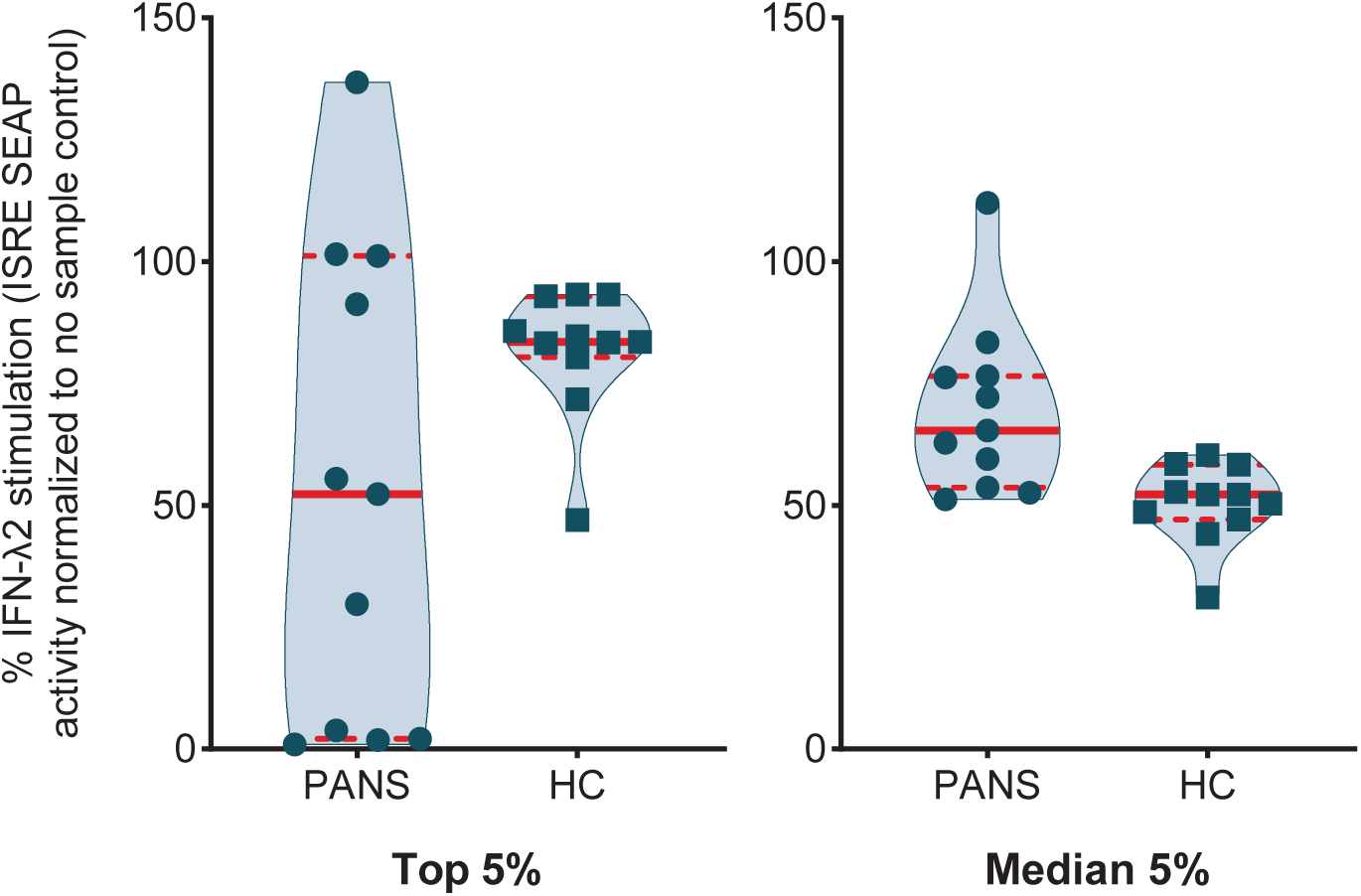
IFN-λ2 neutralization assay results comparing patients and healthy control groups. Samples were selected based on the top or median 5% (n = 11) of MFI values from the IFN-λ autoantibody array screen (from **Fig 2A**). Values reflect relative IFN-λ2 stimulation (at the EC_75_ concentration of 1.4 ng/mL) in the presence of a 1:10 dilution of heat-inactivated plasma after normalization to IFN-λ-only control.

**Supplementary Figure 5.**
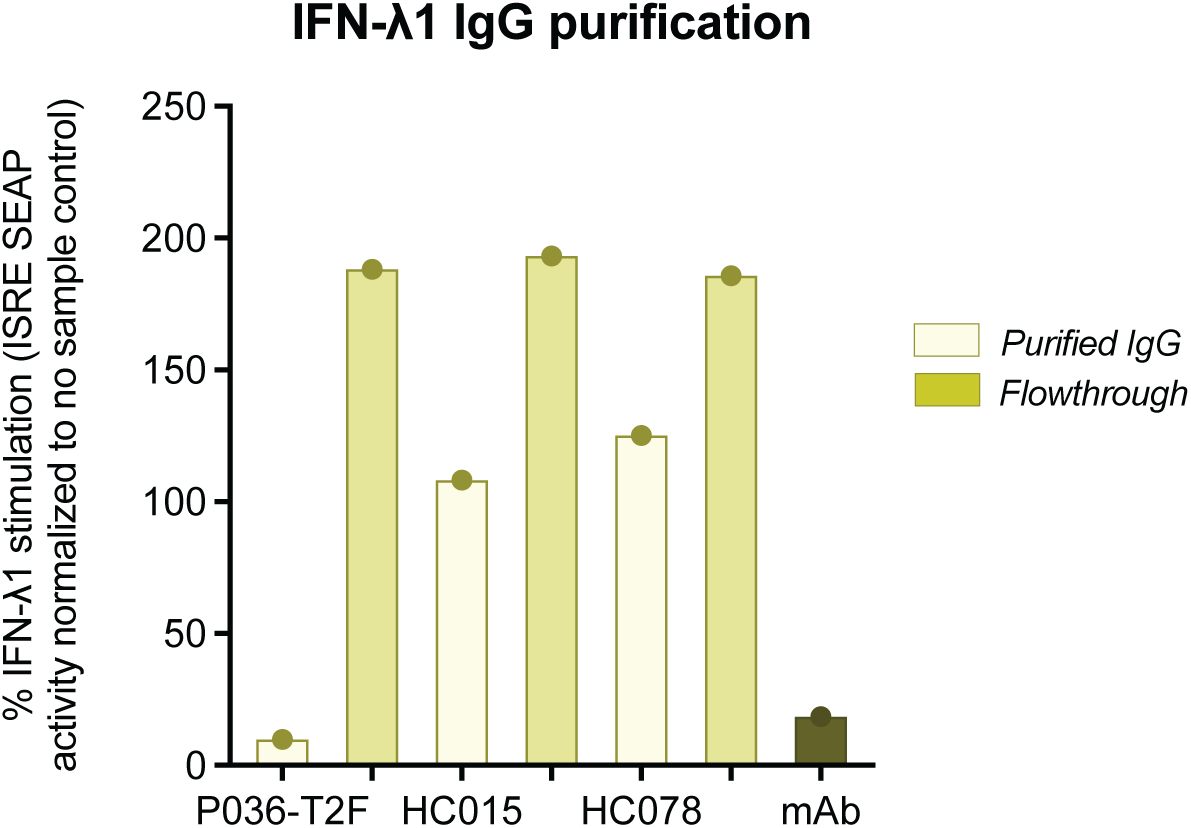
IFN-λ1 neutralization assay comparing purified IgG from patient plasma with IFN-λ1 neutralization activity, and healthy controls without neutralization activity. 20% IgG-depleted flow-through plasma, which was 1:1 diluted during IgG purification, from each purified sample and IFN-λ1 mAbs (5 µg/mL) were included as controls. Samples were incubated with IFN- λ1 (0.5 ng/mL).

## Contributions

X.Y., J.F., P.J.U. and T.R.P. designed experiments and antigen array panels. X.Y., T.R.P., M.K., W.J.K., A.C. and M.J.B. generated antigen arrays. X.Y. performed most sample runs, quality control, cytokine neutralization arrays and data processing. A.C. and M.J.B. performed additional autoantibody screening arrays and interpreted the related data. M.K. and W.J.K. performed and analyzed additional cytokine neutralization arrays. J.F. provided all PANS and HC samples. C.M. assisted with sample collection. J.F., M.M., and B.F. characterized the rheumatologic findings and diseases based on published criteria. M.T., M.S., Y.X, and P.T classified patients based on PANS diagnosis. J.F., N.S. and T.R.P. generated clinical data tables. X.Y., T.R.P. and P.J.U. analyzed and interpreted array data and generated figures. X.Y., T.R.P. J.F. and P.J.U. wrote the paper.

## Acknowledgements

We are grateful for our patients and families who understand treatment limitations and continue to lend their time and cooperation to research participation. We would also like to thank the Stanford IBH Clinic Team, current and former members of our IBH research staff, and our collaborating physicians.

